# Consanguinity, Inbreeding Coefficient, Infant Mortality and congenital anomalies evaluation in the population of Faisalabad

**DOI:** 10.64898/2026.02.01.26345314

**Authors:** Saba Khalid, Mudassar Hassan

## Abstract

**Background:** Consanguineous unions are defined as the matrimony between individuals who are blood relatives. Researchers in all over the world worked on this issue and they checked the ratio of prevalence and effects of consanguinity in different regions of world. This research was conducted in the District Faisalabad, upper Punjab.

**Objective:** To find rate of consanguinity, coefficient of inbreeding (*F*) and its impacts.

**Methods:** The data was collected from six tehsils of District Faisalabad by interviewing the subjects. The data collected within the time span of six months. Total of 2366 subjects were interviewed after their consent approval.

**Results:** The rate of consanguinity was noted 41.83% with 0.03053 coefficient of inbreeding. High rate of consanguinity (23.36%) was noted among first cousins. The distantly related and not related unions were 35.64% and 22.56% respectively. The rate of consanguineous unions in six tehsils ranged from 33.99% in Jaranwala to 53.85% in Tandlianwala. Consanguineous marriages were noted high in Punjabi speaking subjects, in housewives, in reciprocal marital types, in grand-parents and one couple family types and Rajpoot castes. There was found no significant differences of consanguinity in rural and urban areas. The rate of still births was noted high (82.25%) in consanguineous unions while neonatal, post neonatal and child mortality was low such less as 6.45%, 8.06% and 3.22% respectively. The prenatal mortality was noted slightly high 44.94% in consanguineous unions as compared to non-consanguineous unions. The congenital malformation rate was 6.29% in all marital unions but this rate was high (59.06%) in consanguineous unions as compared to non-consanguineous unions (40.93%). This is a pilot study to analyze the potential of inbreeding coefficient (*F*) in the District Faisalabad.

## Introduction

Consanguineous marriages are defined as the relationship between two people who shared same blood line and genetics either from their mother or father and may be from both sides if their parents are also cousins. The marriages which occur as a result of consanguineous relationship enhance the ratio of having a same gene inherited from a mutual forefather (Khan *et al*., 2016). The occurrence of consanguinity differs in various regions of world according to culture, religion, society values and nation (Tadmouri *et al*., 2009). In thousand million people 20% to 50% marriages are consanguineous (Bittles *et al*., 2010). The people who are second cousin or related ones are 10.4% of 6.7 billion global populations (Bittles *et al*.,2010). A demographic survey of 1990 to 1991 showed the rate of consanguinity in Pakistan around 63% and out of which 80 % were first cousins (Sathar *et al*., 1991). The consanguineous marriage frequency was found in the city of Jhelum 44.3%, in Gujrat 48.50%while in Sialkot this ratio varied to 52%and in Sahiwal varied up to 56.1% (Bittles *et al*., 1993), 56.72% in Sargodha (Hina *et al*., 2014), and 62% recorded in Bhimer district, Azad jammu and Kashmir (Jabeen *et al*., 2014). Consanguinity is important because of its impacts directly affecting human health. In first cousin marriages, the rate of hereditary material distribution is four times higher as compare to second cousins (Vanagatie *et al*., 2014). Consanguinity increases the chances of passing damage recessive allele innate from a common ancestor and as a result, the genetic diseases pass from one generation to next and becomes dominant (Masood *et al*., 2011). It is said consanguineous marriages are significant reason behind infant mortality and morbidity because of defective genes (Guo, 1993). According to one study conducted in Pakistan, the rate of first cousin marriages is 1.18 times more as compare to other marriages types which result death of children (Shah *et al*., 1998). The rate of post neonatal mortality and childhood morbidity is three fold in case of consanguinity while another study reports that 50-60% morbidity and mortality eradicated in Pakistan if consanguineous marriages not occur (Bundey and Alam, 1993). In some countries due to the side effects of cousin marriages, consanguinity is ban such as Korea (Ottenheimer, 1996). Women who are married with their cousin are less educated, lower in age at marriage time and have a more reserved perspective to using artificial methods to prevent pregnancy which result short birth intervals, increase in number of childbirths, persistence of giving births at higher ages and many more other things (Basu,1975; Khlat, 1988; Bittles *et al*., 1993). In some states of America and in china the government making strict laws on cousin marriages. Some Churches also banned the consanguineous marriages (Jibril *et al*., 2014). This is the need of time to aware and educates the people on the side effects of consanguinity to avoid its harsh effects in next generation. Media, Newspaper and internet are good sources for awareness of people. To reduce the side effects of consanguinity governments should have established laws and educational and awareness programs.

## Material and Methods

### Location

Faisalabad district is situated in the North-East of Province Punjab, Pakistan (co-ordinates 31°25′0″N 73°5′28″E). Faisalabad lies 184 meters from sea level. In the era of British India, this city was one of most developed cities. Faisalabad city is surrounded by Chiniot and Sheikhupura from north side, Sahiwal and Sheikhupura from east side, Sahiwal and Toba Tek Singh from south side and Jhang from west. The city Faisalabad is situated in flat landform of Himalayan foothills and in the Indian subcontinent central core. The weather of Faisalabad varied from warm in summer to cold in winter. The majority of population of this city is Muslim while minorities also exist. The old name of Faisalabad district was Lyallpur. This city covers an area of 5,856 Km2. Faisalabad got attention because of its industries and distribution centers. This city is the hub of province Punjab because it connects this region with roads, rails and airports. This is the reason, which makes it the Manchester of Pakistan. Twenty percent gross domestic production of Punjab comes from Faisalabad City. Faisalabad has six tehsils; Faisalabad city, Faisalabad sadder, Chak jhumra, Sammundri, Tandlianwala and Jaranwala.

### Data Collection

The data collected within the time span of six months from March 2017 to September 2017. The data was collected from six tehsils of District Faisalabad (Figure 1) by door to door visits, relatives, known ones and from patients of local hospitals and private clinics. The total data of 2366 subjects was collected after taking consent approval. The subjects included in this research were randomly selected married females and all were Pakistani nationals.

**Figure 1.**
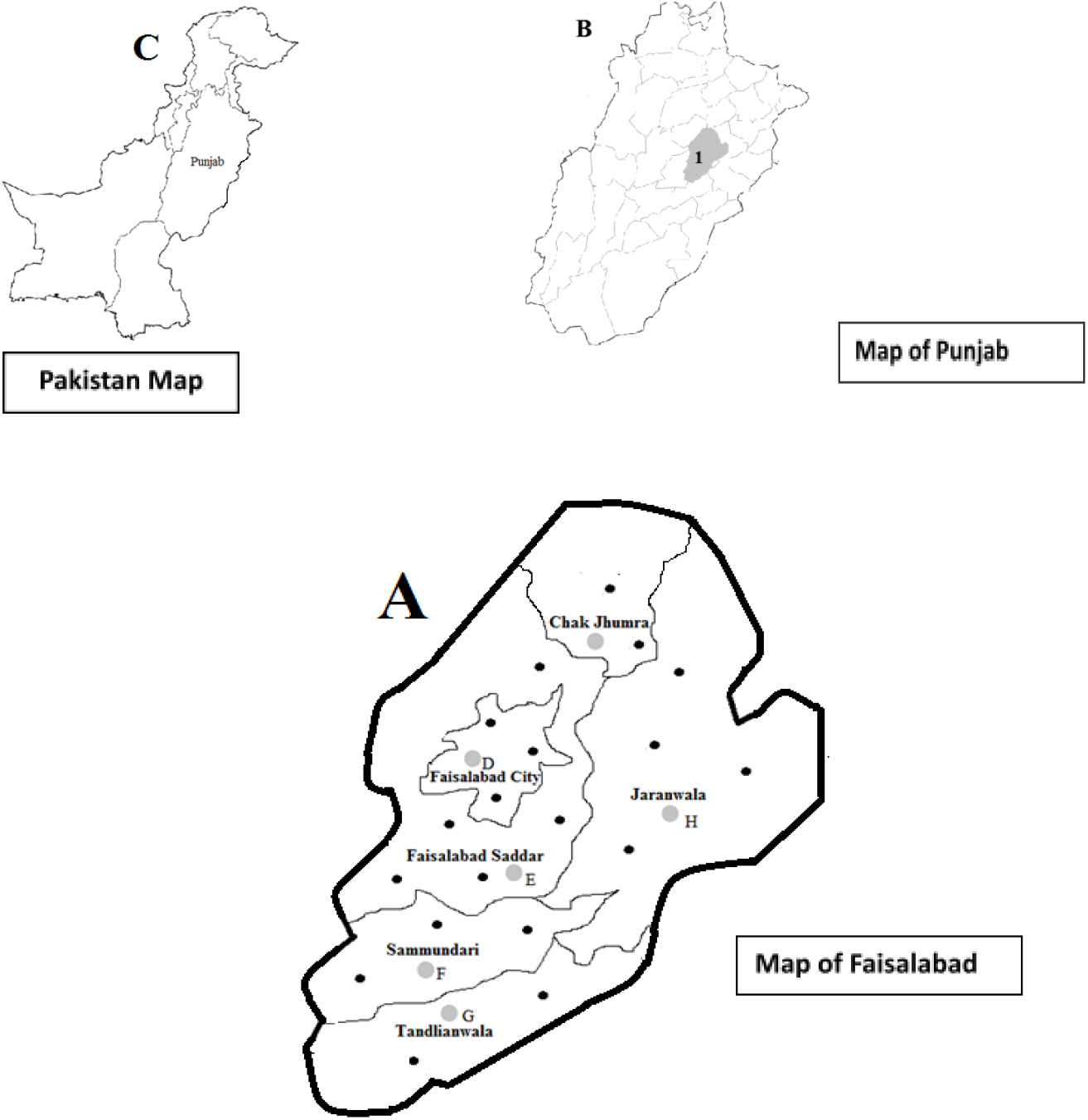
Map of Faisalabad district (A) superimposed on a map of Punjab (B) and Pakistan (C). There were 23 sampling sites in District Faisalabad, depicted as black dots (A).

### Methodology

A designed questionnaire was used for collection of information, which can be define as a device for getting answers to questions by using a form which respondent fills in itself. In questionnaire the questions related to demographic profile of subject, subject parents and spouse (i.e., origin, linguistics, caste, education, age), parents and subject marriage record (i.e., marriage year, marriage type, family type) and disease information. Data on marital union types and socio-demographic attributes of the respondent were acquired. Self-identified ‘occupation’ and ‘economic status’ were recorded. Three family/household types were recognized, i.e. ‘nuclear’, ‘more than one couple’ and ‘extended’. In nuclear families, one couple and their children were living as a unit. In ‘more than one couple’ households, there were two brothers with their wives and children dwelling in the same house. ‘Extended’ households had three-generation families. The data was recorded on marriage arrangements, i.e. arranged, reciprocal and self-arranged/arranged-love marriages. “Arranged marriages” were those in which parents of the subject made the decision about the marriage partner. “Reciprocal marriages” or watta-satta were arranged marriages that involved two exchanged marriages in two families, and ‘self-arranged or arranged-love marriages’ were those in which the subject identified the marriage partner and the marriage went ahead with the consent of both families.

## Results

The total data of 2366 subjects was collected from randomly selected married females whose ages range from 16-86. The rate of consanguineous marriages which noted high in this area was 41.43% while non consanguineous unions noted 58.19%The inbreeding coefficient *F* evaluated 0.03053 in District Faisalabad. The rate of distantly related unions noted 35.63% out of all marital unions. In consanguineous unions rate of first cousin marriages was higher 26.36% as compare to other cousin types. The non-related marital unions noted 22.56% (Table 1).

**Table 1:** Tehsil Wise Distribution of Marital Unions Types in six tehsils of District Faisalabad.

| Tehsil | Consanguineous unions |  |  |  | Non-Consanguineous Unions |  |  | All |
| --- | --- | --- | --- | --- | --- | --- | --- | --- |
|  | Double First Cousin | First Cousin | First cousin once removed | Second Cousin | Second Cousin once removed | Distantly Related Bradri | Non Related |  |
| **Faisalabad City** | 103<br>(8.45%) | 341<br>(27.97%) | 26<br>(2.13%) | 41<br>(3.36%) | 42<br>(3.44%) | 476<br>(39.05%) | 190<br>(15.59%) | 1219<br>(51.52%) |
| **Faisalabad Saddar** | 67<br>(22.56%) | 34<br>(5.46%) | 3<br>(1.01%) | 11<br>(3.70%) | 2<br>(0.67%) | 55<br>(18.52%) | 125<br>(42.08%) | 297<br>(8.70%) |
| **Sammundari** | 18<br>(8.74%) | 74<br>(35.92%) | 9<br>(4.37%) | 2<br>(0.971%) | 3<br>(1.46%) | 72<br>(34.95%) | 28<br>(13.59%) | 206<br>(12.56%) |
| **Jaranwala** | 10<br>(3.27%) | 78<br>(25.49%) | 7<br>(2.29%) | 9<br>(2.94%) | 7<br>(2.29%) | 99<br>(32.35%) | 96<br>(31.37%) | 306<br>(6.60%) |
| **Chak Jhumra** | 5<br>(3.21%) | 49<br>(16.01%) | 4<br>(1.31%) | — | 1<br>(0.641%) | 34<br>(21.8%) | 63<br>(40.38%) | 156<br>(12.93%) |
| **Tandlianwala** | 36<br>(19.78%) | 47<br>(25.82%) | 12<br>(6.59%) | 3<br>(1.65%) | 7<br>(3.85%) | 45<br>(24.72%) | 32<br>(17.58%) | 182<br>(7.69%) |
| **Percentages** | 10.1 | 26.36 | 2.57 | 2.78 | 2.62 | 33.01 | 22.56 | 2366 |
| **Total** | 239 | 623 | 61 | 66 | 62 | 781 | 534 |  |

In six tehsils of District Faisalabad consanguineous unions ranged from 33.99% Jaranwalato 53.85% Tandlianwala.(Table 2). The consanguinity found statistically significant in six tehsils of District Faisalabad in comparison of non-consanguineous unions (p = 0.0001). The rates of consanguineous marriages were noted high in age intervals between 21-25. The consanguineous unions with respect to age intervals increased from older ages to younger ages (Table 2). According to the origin, urban subjects were higher 66.78%. The rates of consanguineous unions were noted slightly higher in rural areas. No significant differences in the rates of consanguinity were found among rural and urban population and the coefficient of inbreeding was approximately similar in both areas. Consanguineous unions noted higher in Punjabi and Urdu speaking subjects with the rates of 42.45% and 38.16% respectively (Table 2). With respect to subject’s family types prominent consanguineous unions noted in more than one couple with the rate of 53.46% with 0.04085 coefficients of inbreeding and statistically significant p= 0.0001. According to marriage arrangement, the highest rate of 65.6% consanguinity noted in reciprocal marital union with significant value of p= 0.0001. The variety of castes groups noted in the subjects of district Faisalabad. The high rate of consanguinity noted in Rajpoot caste with 47.83% and 0.03545 coefficient of inbreeding. In all castes consanguineous unions fluctuated from 32.36% to 47.83% (Table 2). The subjects who were belonging to office jobs occupation had high rate of consanguinity with 65.90% (Table 2). Consanguineous unions were studied according to the occupation of subject’s spouse occupation. The comparison revealed high rate of 60.86% consanguinity in those subject’s spouses who were landlords (Table 2). In 41.81% subject cousin marriage 60.94% parents were consanguineous which was comparatively high with respect to other marital unions (Table 3). The subject and spouse literacy level compared to analyze the difference between their literacy. In 12.43% illiterate subjects, 40.48% of spouses were illiterate and 59.52% literate while out of 87.57% literate subjects, 4.10% spouses were recorded illiterate and 95.90% literate. The analysis showed spouse was highly literate as compare to female subjects. The mean difference analysis of subject and spouse showed that spouse age was comparatively high as compare to female subjects. In cousin marriages the number of pregnancies were high i.e. 42.70% while in distantly related marriages 34.02% and in not related 23.28% which revealed the high rate of fertility in consanguineous unions as compare to other marital unions. The prenatal mortality in consanguineous unions was on slight increased level 44.94% with respect to distantly related marriages 43.20% and in not related marriages the rate was 34%. The rate of still births was high in consanguineous marriages while neonatal, postnatal and child mortality was noted high in distantly related marital union (Table 3). The rate of congenital anomalies was comparatively high in consanguineous unions as compare to non-consanguineous unions. Different types of congenital anomalies noted such as diabetes, limb disorders, microcephaly, albinism, autism, and congenital heart disease, Muscular dystrophy, Rickets, Haemophillia, Cystic fibrosis, cleft lip, cleft palate.

**Table 2:**
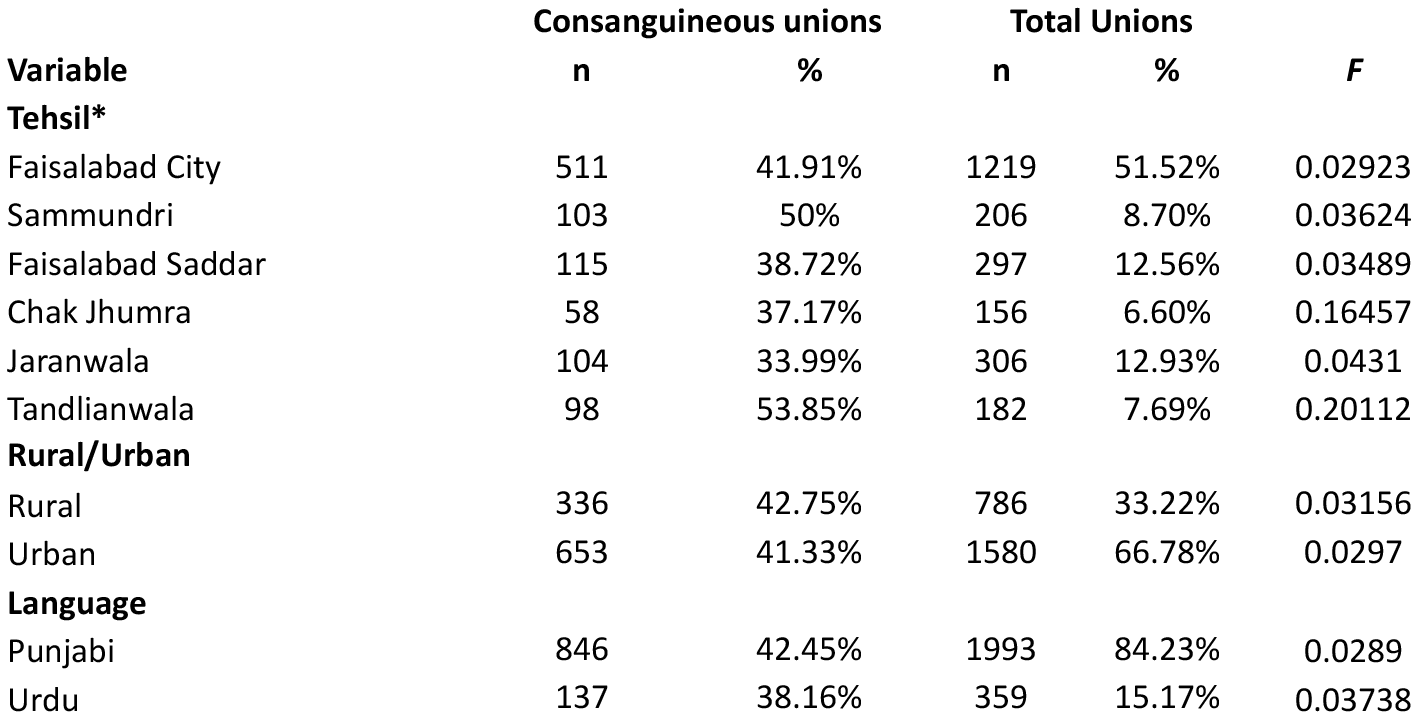
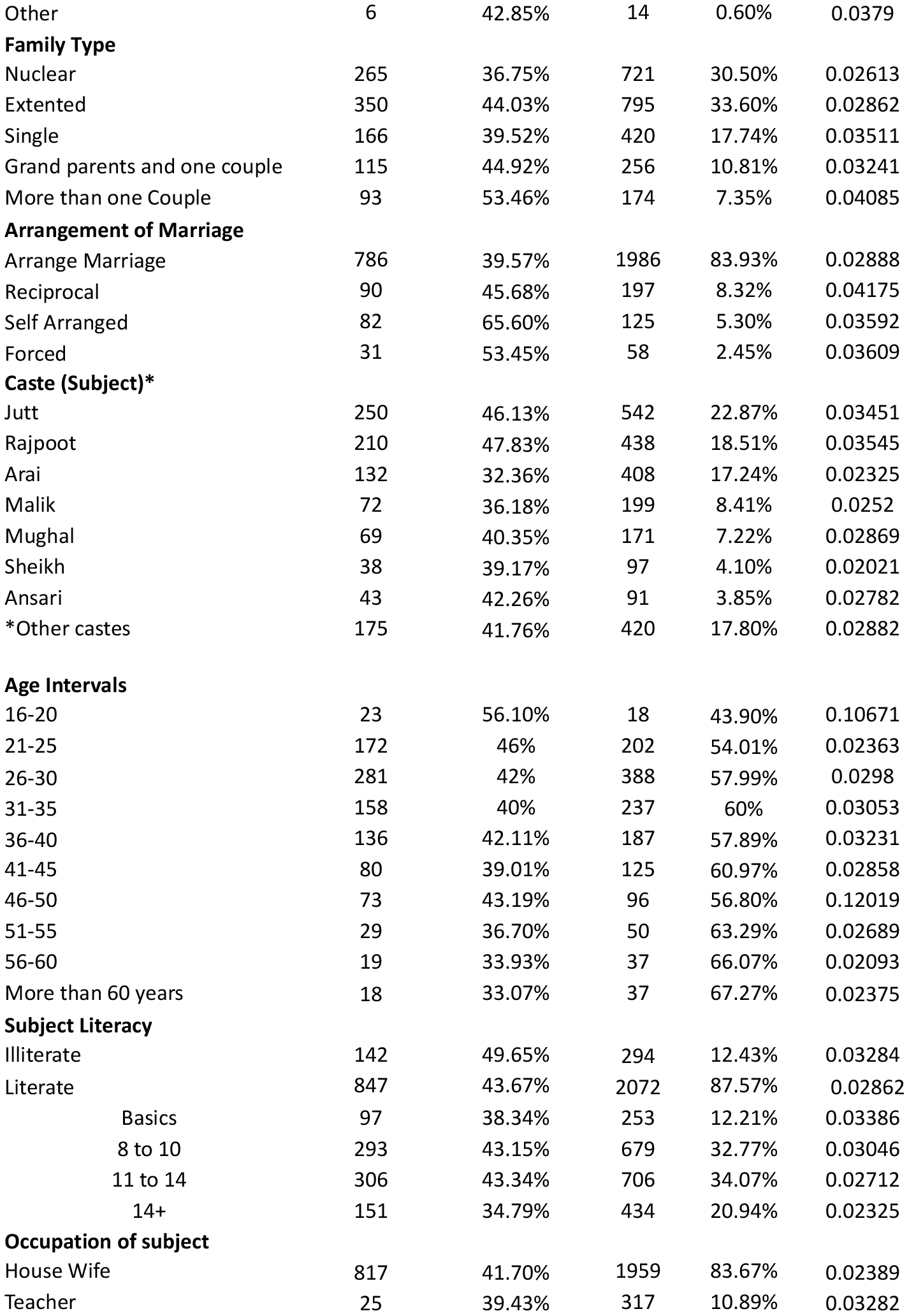
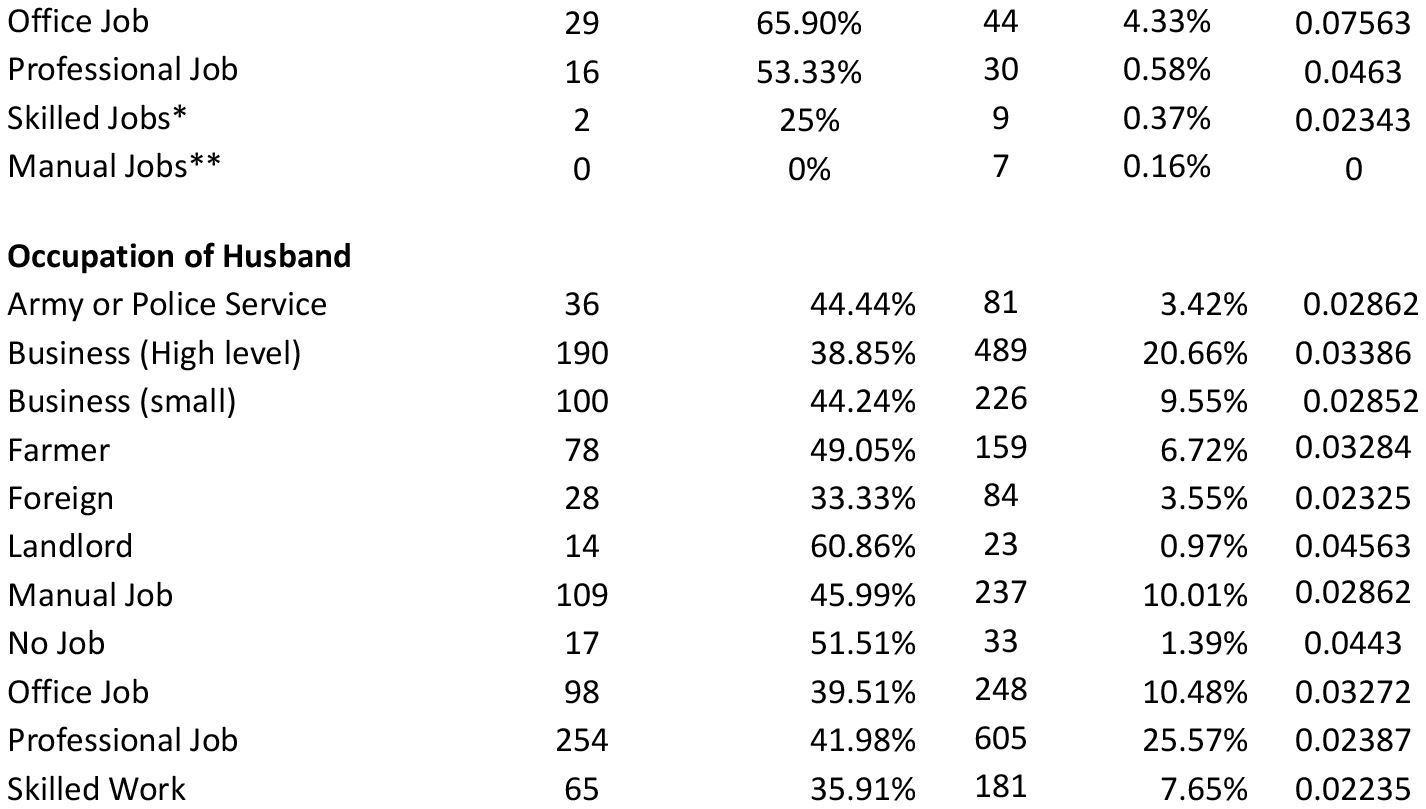
Distribution of consanguineous and total marital union rates and inbreeding coefficients (F) by socio-demographic variables, Faisalabad district, Pakistan.

| Variable | Consanguineous unions |  | Total Unions |  | F |
| --- | --- | --- | --- | --- | --- |
|  | n | % | n | % |  |
| **Tehsil\*** |  |  |  |  |  |
| Faisalabad City | 511 | 41.91% | 1219 | 51.52% | 0.02923 |
| Sammundri | 103 | 50% | 206 | 8.70% | 0.03624 |
| Faisalabad Saddar | 115 | 38.72% | 297 | 12.56% | 0.03489 |
| Chak Jhumra | 58 | 37.17% | 156 | 6.60% | 0.16457 |
| Jaranwala | 104 | 33.99% | 306 | 12.93% | 0.0431 |
| Tandlianwala | 98 | 53.85% | 182 | 7.69% | 0.20112 |
| **Rural/Urban** |  |  |  |  |  |
| Rural | 336 | 42.75% | 786 | 33.22% | 0.03156 |
| Urban | 653 | 41.33% | 1580 | 66.78% | 0.0297 |
| **Language** |  |  |  |  |  |
| Punjabi | 846 | 42.45% | 1993 | 84.23% | 0.0289 |
| Urdu | 137 | 38.16% | 359 | 15.17% | 0.03738 |
| Other | 6 | 42.85% | 14 | 0.60% | 0.0379 |
| **Family Type** |  |  |  |  |  |
| Nuclear | 265 | 36.75% | 721 | 30.50% | 0.02613 |
| Extented | 350 | 44.03% | 795 | 33.60% | 0.02862 |
| Single | 166 | 39.52% | 420 | 17.74% | 0.03511 |
| Grand parents and one couple | 115 | 44.92% | 256 | 10.81% | 0.03241 |
| More than one Couple | 93 | 53.46% | 174 | 7.35% | 0.04085 |
| **Arrangement of Marriage** |  |  |  |  |  |
| Arrange Marriage | 786 | 39.57% | 1986 | 83.93% | 0.02888 |
| Reciprocal | 90 | 45.68% | 197 | 8.32% | 0.04175 |
| Self Arranged | 82 | 65.60% | 125 | 5.30% | 0.03592 |
| Forced | 31 | 53.45% | 58 | 2.45% | 0.03609 |
| **Caste (Subject)\*** |  |  |  |  |  |
| Jutt | 250 | 46.13% | 542 | 22.87% | 0.03451 |
| Rajpoot | 210 | 47.83% | 438 | 18.51% | 0.03545 |
| Arai | 132 | 32.36% | 408 | 17.24% | 0.02325 |
| Malik | 72 | 36.18% | 199 | 8.41% | 0.0252 |
| Mughal | 69 | 40.35% | 171 | 7.22% | 0.02869 |
| Sheikh | 38 | 39.17% | 97 | 4.10% | 0.02021 |
| Ansari | 43 | 42.26% | 91 | 3.85% | 0.02782 |
| *Other castes | 175 | 41.76% | 420 | 17.80% | 0.02882 |
| **Age Intervals** |  |  |  |  |  |
| 16-20 | 23 | 56.10% | 18 | 43.90% | 0.10671 |
| 21-25 | 172 | 46% | 202 | 54.01% | 0.02363 |
| 26-30 | 281 | 42% | 388 | 57.99% | 0.0298 |
| 31-35 | 158 | 40% | 237 | 60% | 0.03053 |
| 36-40 | 136 | 42.11% | 187 | 57.89% | 0.03231 |
| 41-45 | 80 | 39.01% | 125 | 60.97% | 0.02858 |
| 46-50 | 73 | 43.19% | 96 | 56.80% | 0.12019 |
| 51-55 | 29 | 36.70% | 50 | 63.29% | 0.02689 |
| 56-60 | 19 | 33.93% | 37 | 66.07% | 0.02093 |
| More than 60 years | 18 | 33.07% | 37 | 67.27% | 0.02375 |
| **Subject Literacy** |  |  |  |  |  |
| Illiterate | 142 | 49.65% | 294 | 12.43% | 0.03284 |
| Literate | 847 | 43.67% | 2072 | 87.57% | 0.02862 |
| Basics | 97 | 38.34% | 253 | 12.21% | 0.03386 |
| 8 to 10 | 293 | 43.15% | 679 | 32.77% | 0.03046 |
| 11 to 14 | 306 | 43.34% | 706 | 34.07% | 0.02712 |
| 14+ | 151 | 34.79% | 434 | 20.94% | 0.02325 |
| **Occupation of subject** |  |  |  |  |  |
| House Wife | 817 | 41.70% | 1959 | 83.67% | 0.02389 |
| Teacher | 25 | 39.43% | 317 | 10.89% | 0.03282 |
| Office Job | 29 | 65.90% | 44 | 4.33% | 0.07563 |
| Professional Job | 16 | 53.33% | 30 | 0.58% | 0.0463 |
| Skilled Jobs* | 2 | 25% | 9 | 0.37% | 0.02343 |
| Manual Jobs** | 0 | 0% | 7 | 0.16% | 0 |
| Occupation of Husband |  |  |  |  |  |
| Army or Police Service | 36 | 44.44% | 81 | 3.42% | 0.02862 |
| Business (High level) | 190 | 38.85% | 489 | 20.66% | 0.03386 |
| Business (small) | 100 | 44.24% | 226 | 9.55% | 0.02852 |
| Farmer | 78 | 49.05% | 159 | 6.72% | 0.03284 |
| Foreign | 28 | 33.33% | 84 | 3.55% | 0.02325 |
| Landlord | 14 | 60.86% | 23 | 0.97% | 0.04563 |
| Manual Job | 109 | 45.99% | 237 | 10.01% | 0.02862 |
| No Job | 17 | 51.51% | 33 | 1.39% | 0.0443 |
| Office Job | 98 | 39.51% | 248 | 10.48% | 0.03272 |
| Professional Job | 254 | 41.98% | 605 | 25.57% | 0.02387 |
| Skilled Work | 65 | 35.91% | 181 | 7.65% | 0.02235 |

**Table 3:**
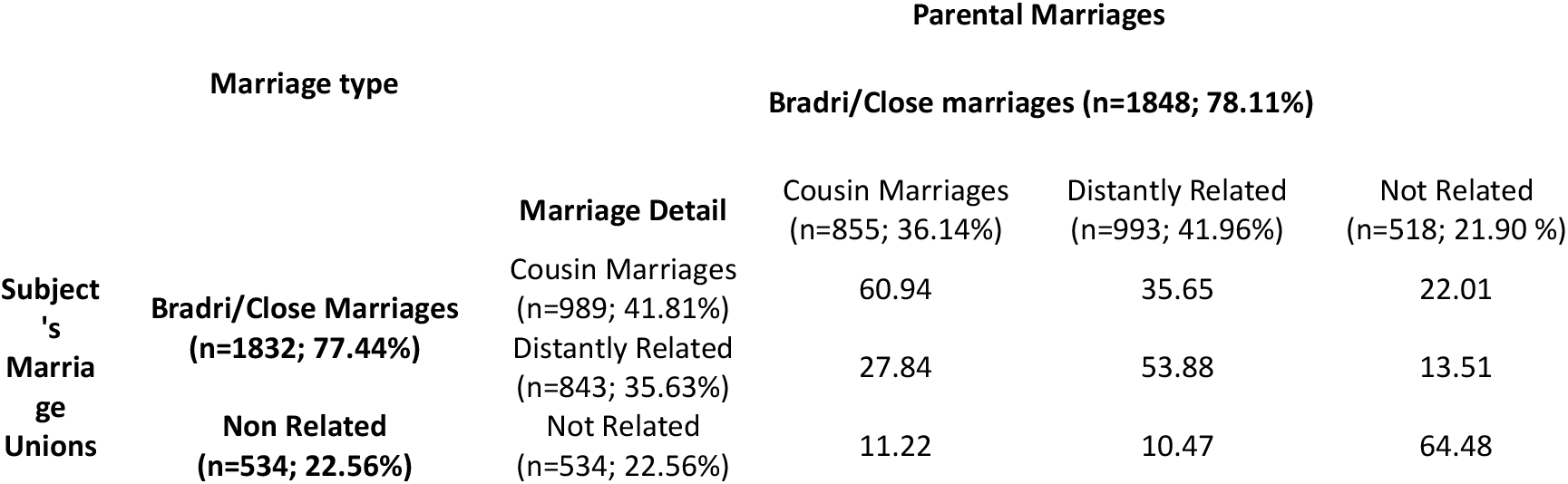
Comparison of subject and parental marital unions.

**Table 4:** Pregnancy outcomes and Consanguineous Unions.

|  | Live Births | Prenatal Mortality | Postnatal Mortality |
| --- | --- | --- | --- |
| **Cousin Marriages (n=989; 41.81%)** | 2475<br>(42.32%) | 129<br>(44.94%) | 62<br>(44.92%) |
| **Distantly Related (n=843; 35.63%)** | 1967<br>(33.63%) | 124<br>(43.20%) | 56<br>(40.58%) |
| **Not Related (n=534; 22.56%)** | 1406<br>(24.04%) | 34 (11.87%) | 20<br>(14.49%) |
| **Total** | 5848 | 287 | 138 |

## Discussion

The study which was conducted in six tehsils of district Faisalabad to analyze the prevalence of consanguinity, coefficient of inbreeding and its effects revealed results which are never recorded in this area before. According to this study, the rate of consanguinity in district Faisalabad was 44.43%. The analysis of the distribution of marital unions in District Faisalabad with other recent researches which conducted on other places depict that the rate of consanguinity which observed in District Faisalabad was comparatively low with respect to District Sargodha (56.72%), Rahim yar khan (58.46%), District Kashmore (68.70%) and (62%) from District Bhimer, Azad Jammu and Kashmir (Hina *et al*., 2014; Riaz *et al*., 2016; Khan *et al*., 2015; Noonari *et al*., 2018) and higher in contrast to Bajaur Agency (22.34%) while coefficient of inbreeding 0.03053 of district Faisalabad has resemled with Bajaur Agency (Ahmad *et al*., 2016). The study which conducted in district Faisalabad cleared that first cousin marriage was most prevalent as compare to other consanguineous unions (Hussain *et al*., 1998). In six tehsils of Faisalabad the rate of consanguinity ranged from 36.27% in Jaranwala to 57.69% in Tandlianwala while in six tehsils of District Sargodha this rate varied 50.38% in Bhalwal to 62.88% Sillanwali which was relatively high as compared to district Faisalabad. The rate of First cousin marriages out of total consanguineous unions were 59.27% while in Rahim yar khan and in Sargodha district the first cousin rate was 52% and 49.11% which was less as compare to District Faisalabad (Riaz *et al*., 2016; Hina *et al*., 2014). It was said 80% marriages in Pakistan noted among first cousin out of 60% total consanguineous unions (Perveen *et al*., 2012). In Pakistan it is so common to marry with their cousin and consanguinity broadly accomplished at society level. The rate of consanguinity in parents of subjects were noted 39.48% which was comparatively low as compare to Rahim Yar Khan where parental consanguinity was 48.5% (Riaz *et al*., 2016). The comparison of subject and parents consanguinity in District Faisalabad showed that the parental consanguinity was 64.45% out of subjects 44.43% consanguineous unions while in Bajaur Agency in 5.2% parental consanguineous marriage 36% subject was consanguineous too (Ahmad *et al*., 2014). This is the trend to marry within the family which prevailed from generation to generation ((Bener A *et al*., 1996).

The close marriages was 42.75% in rural and 41.33% in urban. There was no prominent difference of consanguineous unions within rural and urban area.These results favored the results of Hina *et al*., 2014; Ahmad *et al*., 2016; Jabeen *et al*., 2014 studies which conducted in District Sargodha, Bajaur Agency and Bhimber district and Azad Jammu and Kashmir where consanguinity not significant with area. It was thought that decreasing rates of consanguineous unions in rural areas may be because of the increase shift rate of population from rural to urban areas (Ahmad *et al*., 2016). One more reason of the declining rate was the evolution in the past household trends toward modern trends (Arif *et al*., 2009).

Language varied with respect to regions however different language spoken in the Pakistan. So in this study of District Faisalabad high rate of consanguinity noted in Punjabi speaking subjects just like in District Sargodha (Hina *et al*., 2014) where consanguinity rate prominent in Punjabi speaking people and contrast to Rahim Yar Khan where consanguinity was noted high in Siraki language (Riaz *et al*., 2016). The demographic features varied from area to area so consanguinity depends upon the features of this area. The Punjabi speaking subjects prefer to marry with the Punjabi speaking spouse in Province Punjab but in Province KPK the mostly speaking language was Pashtu so here Pashtu speaking prefer to marry whom who speak Pashtu just like in Khan *et al*., 2015 study which conducted in Buner, Khyber Pakhtunkhwa where consanguinity noted in Pashtuns.

The estimates of consanguineous unions in different age intervals showed high rate of consanguinity 56.09% in 16 to 25 years of age intervals which resembles to Ahmad *et al*., 2016 study conducted in Bajaur where consanguinity noted higher in lower age intervals and in contrast to Rahim Yar Khan where consanguinity increased from lower to older ages (Riaz *et al*., 2016). In District Faisalabad the consanguinity rate declined in literate subjects while in illiterate subjects there was a prominent high rate of consanguinity which favored the Hina *et al*., 2014; Ahmad *et al*., 2016 and Riaz *et al*., 2014 results of District Sargodha, Bajaur Agency and District Rahim yar Khan but opposes Noonari *et al*., 2018 results where no prominent difference found between consanguinity and literacy. It is said that as literacy level of females subject’s increase they reached into older ages and chance of consanguinity decreased (Hussain & Bittles, 2004). The subject literacy rate was 87.57% while their spouse literacy rate was 91.37% but in Sargodha district these rates was 29% and 46% for subjects and spouses (Hina *et al*., 2014). In district Faisalabad high consanguinity rate was noted in prominent castes Rajput, Ansari and in Jutt just like in Hina *et al*., 2014 study where consanguinity noted higher in Jutt and Rajput too.

The fertility of subjects in District Faisalabad noted 42.70% in consanguineous unions while in non-consanguineous unions this rate was 23.28% comparatively less as compare to consanguineous unions favored Kerkeni *et al*., 2007 results extracted from Monastir, Tunisia and also Fareed *et al*., 2016 study which conducted in Jammu region. In both of these regions gross fertility rate seen in subjects. It said that in case of cousin marriages couples are younger in age and highly fertile (Kerkeni *et al*., 2007). The still births rate high in consanguineous unions which favored the findings of Shawky *et al*., 2013 who noted 80% consanguinity in still births and neonatal, postnatal and child mortality rate noted lower in district Faisalabad which opposed David *et al*., 2002 findings. It seen that the ratio of prenatal and postnatal mortality was high in Pakistani natives with the rate of 14.6 per 1000 while for others this value varied to 8.4 (Bundey *et al*., 1990). In district Faisalabad district consanguinity rate was noted high in prenatal mortality but not with prominent difference with respect to other marital unions but it favored the findings Hosseni *et al*., 2014. The consanguinity in parents increased the chances of deleterious alleles to pass into next generations cause different type of congenital malformations. The high rate of congenital malformation noted in consanguineous unions as compare to non-consanguineous unions. These results proofed right the Riaz *et al*., 2016 findings which showed that parental consanguinity increase chances of congenital malformation two fold in next generation. The congenital malformations which noted in increased rate was Diabetes, Eye and ears congenital malformation, Spina Bifida, Limb disorder, Rickets, Thallassemia, congenital heart diseases and some others. These disorders because of consanguiniy were found by Iqbal *et al*., 2014 who observed hearing loss, Ain *et al*., 2011 who noted Thalassemia, Maheswari *et al*., 2016 observed Mental retardation, Epilepsy and congenital blindness.

The present study describes the consanguinity parameters in a representative population of District Faisalabad, which provides the useful data on consanguinity and the inbreeding coefficient. The data illustrate a wide variation in consanguinity and inbreeding coefficient across the socio-demographic strata of the district Faisalabad. Furthermore, comparison of Faisalabad with other populations of Punjab indicated substantial regional heterogeneity in the distribution of consanguinity. Further studies in the adjoining areas would be helpful to understand the consanguinity landscape in the region.

## Data Availability

All data produced in the present work are contained in the manuscript

## Acknowledgements

I gratefully acknowledges the HEC, Pakistan for providing financial support of this research project (No 21. 1069/ SRGP/ R&D/HEC/2016).

